# The association of diet and physical inactivity with obesity, diabetes, and hypertension among older adults living in Sierra Leone

**DOI:** 10.1101/2024.10.30.24316415

**Authors:** Tahir Bockarie, Ankit Shanker, Mohamed B. Jalloh, Alhaji M. Kamara, Maria Lisa Odland, Haja Wurie, Rashid Ansumana, Joseph Lamin, Miles Witham, Oyinlola Oyebode, Justine Davies

**Affiliations:** Faculty of Life Sciences & Medicine, School of Life Course & Population Sciences, Department of Women & Children’s Health, King’s College London, London WC2R 2LS, UK; Warwick Medical School, University of Warwick, Coventry, UK; Department of Medicine, McMaster University, 20 Copeland Avenue, Suite C3-117, Hamilton, ON L8L 0A3, Canada; Institute of Global Health, Heidelberg University, Im Neuenheimer Feld 365, Heidelberg, Germany, 69120; Institute of Applied Health Research, College of Medical and Dental Sciences, University of Birmingham, Birmingham B15 2TT, UK; College of Medicine and Allied Health Sciences, University of Sierra Leone, Freetown, Western Area, Sierra Leone; School of Community Health Sciences, Njala University, Bo Campus, Bo, Sierra Leone; Mercy Hospital Research Laboratory, Bo, Sierra Leone; AGE Research Group, NIHR Newcastle Biomedical Research Centre, Newcastle University, Newcastle upon Tyne, UK; Newcastle upon Tyne Hospitals Trust, Newcastle upon Tyne, UK; Centre for Public Health and Policy, Wolfson Institute of Population Health, Queen Mary University of London, London, UK; MRC/Wits Rural Public Health & Health Transitions Research Unit (Agincourt), University of the Witwatersrand, Johannesburg, South Africa; Department for Global Health, Centre for Global Surgery, Stellenbosch University, Stellenbosch, South Africa

**Keywords:** Cardiovascular diseases, Diet, Physical inactivity, Sierra Leone, Public health interventions

## Abstract

**Introduction:** Cardiovascular diseases (CVDs) are primarily driven by modifiable behavioural risk factors and are the leading cause of death globally. In low- and middle-income countries (LMICs), rapid urbanization and changes in behaviour are exacerbating the burden of CVDs but there is limited research into specific associations with CVDs in older adults in low-income countries, such as Sierra Leone. This study examines the association between behavioural risk factors and their physiological sequalae among adults aged 40 and above in Bo District, Sierra Leone.

**Methods:** A cross-sectional study design was used to collect data from 1,978 randomly sampled adults through a household survey. Survey questions were based on the validated WHO STEPs questionnaire. Multivariable logistic regression analysis was performed to determine the associations between behavioural risk factors (such as diet, physical activity, and salt intake) and the presence of hypertension, diabetes, and obesity, adjusting for socio-demographic variables.

**Results:** At least one physiological risk-factor for CVD was present in 43.5% of participants. Hypertension was associated with urban living (OR=1.46, 95% CI [1.41-1.51]), older age (OR for 80+=3.98, 95% CI [3.70-4.28]), insufficient fruit and vegetable intake (OR=1.52, 95% CI [1.46-1.60]), and low physical activity (OR=1.35, 95% CI [1.27-1.43]). Diabetes was associated with urban residence (OR=1.84, 95% CI [1.66-2.05]), older age (OR for 70-79 =3.82, 95% CI [3.28-4.45]), low fruit and vegetable consumption (OR=1.61, 95% CI [1.36-1.90]), high salt intake (OR=1.34, 95% CI [1.21-1.49]), and low physical activity (OR=1.47, 95% CI [1.26-1.71]). Obesity was less likely among males (OR=0.37, 95% CI [0.36-0.38]) and those aged 80+ (OR=0.39, 95% CI [0.35-0.43]).

**Conclusion:** This study highlights the need for evidence for targeted public health interventions in Bo District, Sierra Leone. Promoting healthier lifestyles to reduce poor diet, and physical inactivity, particularly in urban areas, could decrease CVD prevalence.

**Summary:** *What is already known on this topic:*

- Cardiovascular diseases (CVDs) are the leading cause of global mortality, with a disproportionate burden in low- and middle-income countries.
- Modifiable risk factors, including diet and physical inactivity, significantly contribute to CVD prevalence.
- Limited research exists on the specific associations between these risk factors and CVDs in older adults in Sierra Leone.

*What this study adds:*

- This study provides evidence of significant associations between dietary habits, salt intake, physical activity levels, and physiological risk factors for CVD: hypertension, diabetes, and obesity among adults aged 40 and above in Bo District, Sierra Leone.
- It quantifies the increased odds of physiological CVD risk factors associated with inadequate fruit and vegetable intake, high salt consumption, and physical inactivity in this population.
- The study illuminates the links between behavioural and physiological risk factors in the context of rapid urbanisation in Sierra Leone.

*How this study might affect research, practice or policy:*

- These findings will directly inform the development of targeted public health interventions in Sierra Leone, prioritizing the promotion of fruit and vegetable consumption and increased physical activity among adults.
- The study results will drive policy changes to address urbanisation-related lifestyle factors, including initiatives to improve access to healthy foods and create environments conducive to physical activity.
- This research establishes a foundation for future studies on CVD risk factors in Sierra Leone, particularly focusing on age- and gender-specific interventions

## Introduction

Non-communicable diseases (NCDs), including cardiovascular disease (CVD) are the leading causes of mortality worldwide, accounting for 78% of global deaths, constituting a significant public health challenge ^1–4^. In 2019, CVDs alone were responsible for 17.9 million deaths globally, underscoring their significant public health relevance ^5^ ^6^. Low- and middle-income countries (LMICs) face a dual burden of infectious and NCDs, straining healthcare systems and hindering economic development ^11^. Research has highlighted that both LMICs and the vulnerable communities of high-income countries (HIC) share a disproportionate burden of CVDs ^6^. Furthermore, the high prevalence of these risk factors poses a substantial obstacle to achieving Sustainable Development Goal 3, which aims to ensure healthy lives and promote well-being for all at all ages by 2030 ^3^ ^11^.

Drivers of CVDs are largely behavioural, including dietary behaviour, alcohol consumption leading to physiological risk factors, including hypertension, diabetes and obesity ^4 7 8 9^. Understanding the interplay between these behavioural and physiological risk factors in different populations is crucial for developing effective public health strategies aimed at mitigating the prevalence of CVDs.

Although there is growing understanding of behavioural risk factors for CVDs in many LMICs, there is less knowledge about these factors in some of the most deprived countries in sub-Saharan Africa (SSA). This gap is concerning, especially given the rising prevalence of CVDs in these regions amid rapid demographic shifts. Urbanization often leads to lifestyle changes including increased consumption of processed foods and reduced physical activity due to greater reliance on motorized transportation ^12–16^. Studies indicate that urban dwellers in LMICs exhibit dietary patterns characterized by insufficient intake of fruits and vegetables, high consumption of sugar and salt, and increased intake of vegetable oil, meat, and processed foods ^9^ ^10^ ^17–21^ [10, 11, 20-23]. Systematic reviews have highlighted that salt intake in sub-Saharan Africa is alarmingly high, often exceeding recommended levels due to the consumption of processed foods and added salt. Unhealthy diets, characterized by high consumption of processed foods, sugars, and salt, and low intake of fruits and vegetables, are closely linked to the development of hypertension and obesity ^9^ ^10^. Furthermore, research on dietary consumption patterns in sub-Saharan Africa reveals a complex interaction between meat, fruit, and vegetable intake. While meat consumption has increased, often driven by urbanization and higher income levels, fruit and vegetable intake remain critically low. This imbalance in dietary patterns is linked to rising non-communicable diseases and underscores the need for targeted nutritional interventions ^22^.

Although previous studies have explored the association between behavioural and physiological risk factors for CVDs, contemporary evidence from local populations is necessary to drive action by policymakers. Notably, in many low-income countries, comprehensive studies are lacking that investigate the physiological risk factors for CVDs in adults and their association with dietary and physical activity risk factors. Sierra Leone is one such country where insight into the association between behavioural and physiological risk factors of CVDs in Sierra Leone is needed, given the country’s rapid epidemiological transition^23–26^.

This study aims to examine the association between behavioural and physiological risk factors for CVDs among adults aged 40 years and older in Bo District, Sierra Leone. By elucidating these relationships, we seek to provide evidence-based insights to inform targeted interventions, thereby reducing the burden of CVDs in Sierra Leone and similar settings. This research is crucial for informing policy and practice, ultimately contributing to improved health outcomes and the attainment of global health targets.

## Methods

### Study setting

This study was conducted in Bo District, located in the Southern Province of Sierra Leone. Incorporating the country’s second-largest city, Bo District encompasses well-defined rural and urban areas and is like other districts in Sierra Leone, apart from the Western Area, which is predominantly urban. According to the 2015 census, Bo District had a population of 575,478, with 380,307 (66.1%) residing in rural areas and 195,171 (33.9%) living in urban areas, primarily in Bo City. Notably, only 17.4% (100,188) of the population were over 40 years of age ^27^.

### Sampling strategy

A cross-sectional household survey was conducted from September to December 2018. The primary objective was to collect data on CVD risk factors, with a specific focus on diabetes prevalence, which was anticipated to be the lowest among the CVD risk factors. Initially, target sample size was 1,893 participants, calculated to detect a diabetes prevalence of 4% with ±1% precision. To account for potential non-response and missing data, an oversampling rate of 10% was applied, bringing the total target to approximately 2,082 participants. During data collection, we successfully recruited 2,071 individuals.

Participants in this study were individuals aged 40 years and older, selected from both rural and urban areas. The sampling proportions were aligned with the habitation patterns documented in the 2015 population and housing census for Bo District. This alignment ensured that the sample was representative of the population distribution in terms of age and urban-rural residency. Exclusion criteria included confirmed pregnancy and diagnosis with a terminal or incapacitating illness, which could have potentially influenced the study’s outcomes and participant availability.

To accurately reflect the urban-rural population ratio, the study aimed to include 700 urban and 1,300 rural participants. We purposively sampled the only two urban chiefdoms (Bo City and Tikonko). Out of the 24 eligible urban communities within Bo City and Tikonko, seven were selected from a randomly ordered list, with 100 participants recruited in each. For the rural sample, Seven out of the 14 rural chiefdoms were randomly selected using a random ordering method. We aimed to recruit 93 participants from each selected settlement or village. If the target number was not achieved in the initial community, additional communities were selected from the randomly ordered list until the quota was reached.

Data collection in each community followed a systematic approach, ensuring comprehensive coverage and accuracy. Data collectors began at randomly chosen points within each community and sampled every second household along roads or tracks. Each household was permitted to enrol a maximum of two eligible individuals aged. In smaller communities with fewer than 93 eligible individuals, all individuals over 40 years were included in the study. If a selected participant declined to participate, another individual within the household or community was chosen. In cases where no one was home, a message was left with neighbours, and the researchers returned the following day to complete the sampling.

The study’s geographical scope was limited to a 40 km radius from the centre of Bo, ensuring accessibility for researchers and efficient data collection. All chiefdoms and subdistricts in Bo District fell within this radius, allowing for a comprehensive and representative sample of the district’s population.

## Data collection

Data were collected electronically using ODK® (Open Data Kit) software on tablet devices by a team of 15 trained data collectors. The survey was based on the World Health Organization’s STEPwise approach to chronic disease risk factor surveillance (WHO-STEPS).

WHO-STEPS methodology provides a standardized framework for collecting, analysing, and disseminating data on non-communicable diseases (NCDs) and their associated risk factors.

The household survey incorporated all three steps of the WHO-STEPS approach:

1. **Step 1: Questionnaire** - A comprehensive questionnaire was administered, covering sociodemographic characteristics, behavioural and dietary habits, and medical history related to hypertension and diabetes. The demographic section included questions on age, sex, marital status, and education, while behavioural queries addressed fruit and vegetable intake, physical activity levels, salt intake, and smoking habits. The medical history component inquired about past incidences of stroke, heart attack, or angina, as well as previous treatments for raised blood pressure and cholesterol levels. Additionally, the questionnaire included 49 items related to household assets, construction materials, and awareness of CVD risk factors and treatments.
2. **Step 2: Physical Measurements** - Physical measurements were conducted to obtain anthropometric data and blood pressure readings. Height was measured with a measuring stick with participants standing without shoes, backs, hips, and heels against a wall, and looking ahead horizontally. This method was validated using the SECA 213 stadiometer during training and bi-weekly checks. Body weight was recorded using an Omron medical scale, calibrated daily. Body Mass Index (BMI) was calculated as weight in kilograms divided by the square of the height in meters (kg/m²). Blood pressure was measured using an Omron M6 AC LED blood pressure monitor, with three readings taken five minutes apart; the average of the last two readings was recorded. Participants were seated and rested during these measurements.
3. **Step 3: Biochemical Measurements** - Blood samples were obtained in the morning after an 8-hour overnight fast. The Accutrend Plus® point-of-care device (Roche Diagnostics) was utilized to measure fasting capillary glucose concentrations; results were converted to plasma glucose levels using a conversion factor of 1.11. Participants’ fasting status was verified prior to blood sampling, and those who had not fasted were labelled accordingly.

### Demographic Information

Demographic information included geographic area, sex, and age. Geographic area was categorized as either rural or urban, Sex was classified as female or male. Age was recorded both as a continuous variable and presented as a categorical variable, divided into five groups: 40-49 years, 50-59 years, 60-69 years, 70-79 years, and 80+ years.

### Outcome Measures

The study focused on three primary outcome measures: overweight or obesity, hypertension, and diabetes. Overweight or obesity was defined as having a BMI of ≥25 kg/m², while a BMI <25 kg/m² was used as the reference category. Hypertension was defined by a systolic blood pressure (SBP) of ≥140 mmHg or a diastolic blood pressure (DBP) of ≥90 mmHg, or current use of blood pressure-lowering medication. Diabetes was identified by a fasting plasma glucose level of ≥7.0 mmol/L, a random plasma glucose level of ≥11.1 mmol/l, or the use of antidiabetic medications.

### Behavioural risk-factors

Behavioural risk-factors included dietary habits, specifically fruit, vegetable and salt intake, and physical activity levels. Consumption of fruits and vegetables were reported as servings using show cards provided by data collectors. Low consumption was defined as fewer than five servings daily (<5FV), aligning with global dietary recommendations for fruit and vegetable intake. For salt intake, participants, could select from four responses: (1) added salt at the table, (2) added salt during cooking, (3) consumed salty snacks, or (4) did not use salt from any of these sources. Responses were categorised into three groups: those who consumed salt from only one source (either table salt, cooking salt, or salty snacks), those who consumed salt from two or more sources, and those who did not use salt from any of the specified sources.

Physical activity levels were assessed based on self-reported hours of activity per week, in accordance with previous studies^21^. Total hours and minutes of activity across work, travel, and leisure domains were aggregated into two categories: less than 150 minutes per week (inadequate moderate and vigorous physical activity (MVPA)) and 150 minutes or more per week (adequate MVPA). Physical inactivity was assessed by asking participants “Excluding sleeping, how much time do you usually spend sitting or reclining on a typical day?” The total daily hours were then categorised into two groups: less than 3 hours and 3 or more hours. A composite variable was created to capture overall physical activity risk. Participants were categorized into three groups: no risk factors (adequate MVPA and less than 3 hours of daily inactivity), one risk factor (either inadequate MVPA or 3 or more hours of daily inactivity), and two risk factors (both inadequate MVPA and 3 or more hours of daily inactivity).

To evaluate the combined impact of multiple behavioural risk factors, we created a compound variable ranging from 0 to 6, reflecting the number of risk factors each individual exhibited, with risk-factors aligned to the definitions above. The behavioural risk factors considered were: (1) insufficient fruit and vegetable consumption (0 or 1); (2) salt intake from (0 (no sources), 1 (just one source), 2 (for consumption of salt from two or three sources, where the sources are table salt, cooking salt and salty snacks)), (3) low physical activity (0 for adequate MVPA and less than 3 hours of daily inactivity, 1 for either inadequate MVPA or 3 or more hours of daily inactivity, and 2 for having both physical activity risk factors (both inadequate MVPA and 3 or more hours of daily inactivity). Participants were assigned scores based on the cumulative presence of these risk factors. A score of 0 was assigned to individuals with no risk factors, while a score of 6 was assigned to those exhibiting all six risk factors.

### Ethical Considerations

The study protocol was reviewed and approved by the Sierra Leone Ethics and Scientific Review Committee (SLESRC) at the Ministry of Health and Sanitation (MoHS) and King’s College London (reference: HR-17/18–7298). Consent to conduct the study in each area was sought from chiefs in each subdistrict or village prior to data collection. Written informed consent was obtained from all participants, with literate witnesses assisting in cases of illiteracy. Where signatures were not possible, thumbprints were used. Data used in the analysis was anonymized to protect participants’ privacy

### Statistical Analysis

Statistical analyses were performed using the Statistical Package for Social Sciences (SPSS) version 27 (IBM Corp., Armonk, NY, USA). Participants with missing data on lifestyle factors, outcome measures, or covariates were excluded from the analysis. Descriptive statistics, including frequencies (proportions) and means with standard deviations (SD), were used to summarize the data. The data were weighted using probability weights based on age and sex distributions from the 2015 Population and Housing Census for Bo District. A p-value of ≤0.05 was considered statistically significant.

Multivariable adjusted binary logistic regression models were used to analyse the associations between lifestyle factors and clinical outcomes (hypertension, diabetes, and obesity). The models incorporated all behavioural and socio-demographic covariates simultaneously, without using stepwise selection. This approach ensured a comprehensive evaluation of the relationships between behaviours and health outcomes.

## Results

### Summary of population characteristics, cardiovascular conditions, and risk factors

After excluding 93 cases due to missing values, the final sample consisted of 1,978 individuals. The sociodemographic characteristics of the unweighted and weighted study populations are presented in Table 1.

**Table 1:** Socio-demographic characteristics of study population.

| Household Survey Participants |  |  |
| --- | --- | --- |
|  | Unweighted<br>n (%) | Weighted<br>n (%) |
| Location |  |  |
| Rural | 1205 (60.9) | 60739 (62.6) |
| Urban | 773 (39.1) | 36217 (37.4) |
| Sex |  |  |
| Female | 1100 (55.6) | 47766 (49.3) |
| Male | 878 (44.4) | 49190 (50.7) |
| Age Group |  |  |
| 40-49 | 682 (34.5) | 43116 (44.5) |
| 50-59 | 546 (27.6) | 24379 (25.1) |
| 60-69 | 340 (17.2) | 14381 (14.8) |
| 70-79 | 251 (12.7) | 8913 (9.2) |
| 80+ | 159 (8.0) | 6166 (6.4) |
| Education |  |  |
| Some Education | 615 (31.1) | 31731 (32.7) |
| No Education | 1363 (68.9) | 65225 (67.3) |
| Marital Status |  |  |
| Married or Cohabiting | 1355 (68.5) | 70364 (72.6) |
| Not Married | 623 (31.5) | 26591 (27.4) |
| Wealth Quintile |  |  |
| Poorest | 395 (20.0) | 19896 (20.5) |
| Poorer | 397 (20.1) | 19964 (20.6) |
| Middle | 397 (20.1) | 19493 (20.1) |
| Richer | 394 (19.9) | 19093 (19.7) |
| Richest | 395 (20.0) | 18510 (19.1) |

Among the unweighted study participants, there were 878 men (44.4%) and 1,100 women (55.6%). The majority of participants were between 40-49 years old (34.5%), had no formal education (68.9%), and were married or cohabiting (68.5%). The weighted study population characteristics were similar to those of the unweighted population.

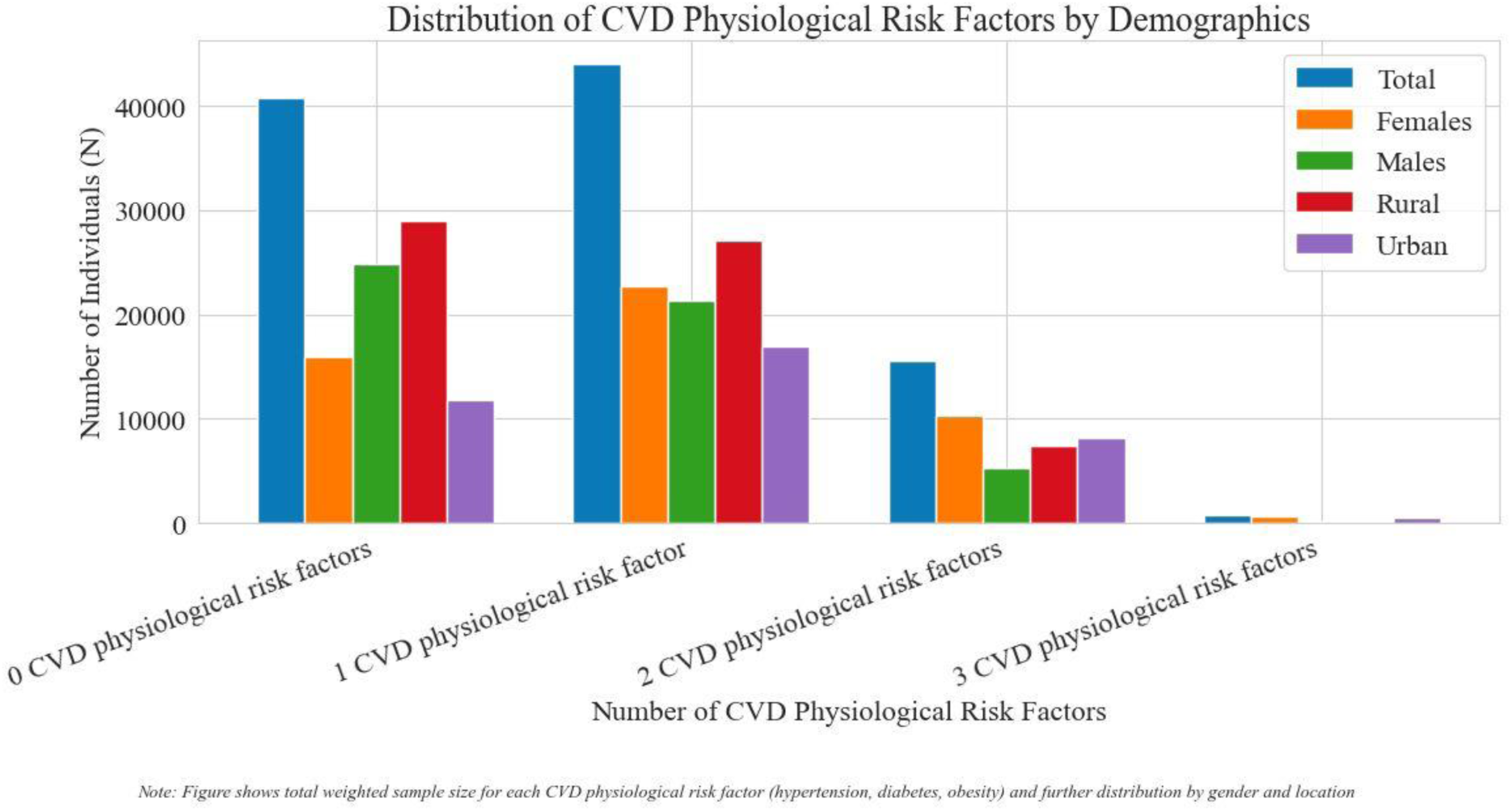

Out of the 1,978 participants, 43.5% had at least one physiological risk factor for CVD, as shown in Table 2. A higher proportion of females (45.9%) compared with males (41.3%) reported having at least one of these conditions. Additionally, fewer rural dwellers (42.6%) had at least one condition compared with urban dwellers (45.0%).

**Table 2.**
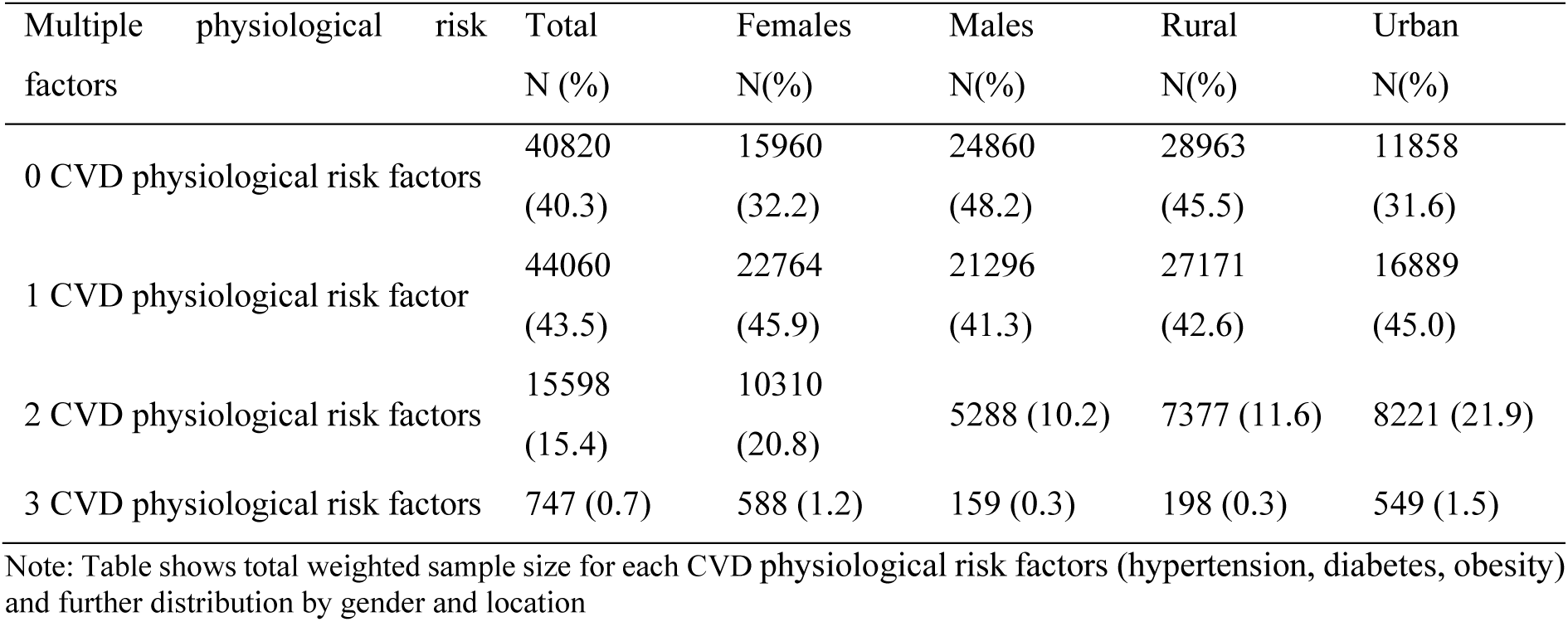
Summary of number of co-occurring physiological risk factor by sex and by location.

| Multiple physiological risk factors | Total<br>N (%) | Females<br>N(%) | Males<br>N(%) | Rural<br>N(%) | Urban<br>N(%) |
| --- | --- | --- | --- | --- | --- |
| 0 CVD physiological risk factors | 40820<br>(40.3) | 15960<br>(32.2) | 24860<br>(48.2) | 28963<br>(45.5) | 11858<br>(31.6) |
| 1 CVD physiological risk factor | 44060<br>(43.5) | 22764<br>(45.9) | 21296<br>(41.3) | 27171<br>(42.6) | 16889<br>(45.0) |
| 2 CVD physiological risk factors | 15598<br>(15.4) | 10310<br>(20.8) | 5288 (10.2) | 7377 (11.6) | 8221 (21.9) |
| 3 CVD physiological risk factors | 747 (0.7) | 588 (1.2) | 159 (0.3) | 198 (0.3) | 549 (1.5) |

Note: Table shows total weighted sample size for each CVD physiological risk factors (hypertension, diabetes, obesity) and further distribution by gender and location

Table 3 shows an increase in the odds of CVD physiological risk factors (Hypertension and Obesity) with an increasing number of behavioural factors; individuals with six behavioural risk factors had a significantly greater risk of hypertension (OR=2.15, 95% CI [1.88-2.44], p<0.001) and overweight/obesity (OR=2.41, 95% CI [2.09-2.78], p<0.001) compared with those in the reference category. For hypertension and obesity, there is a clear dose-response relationship.

**Table 3.**
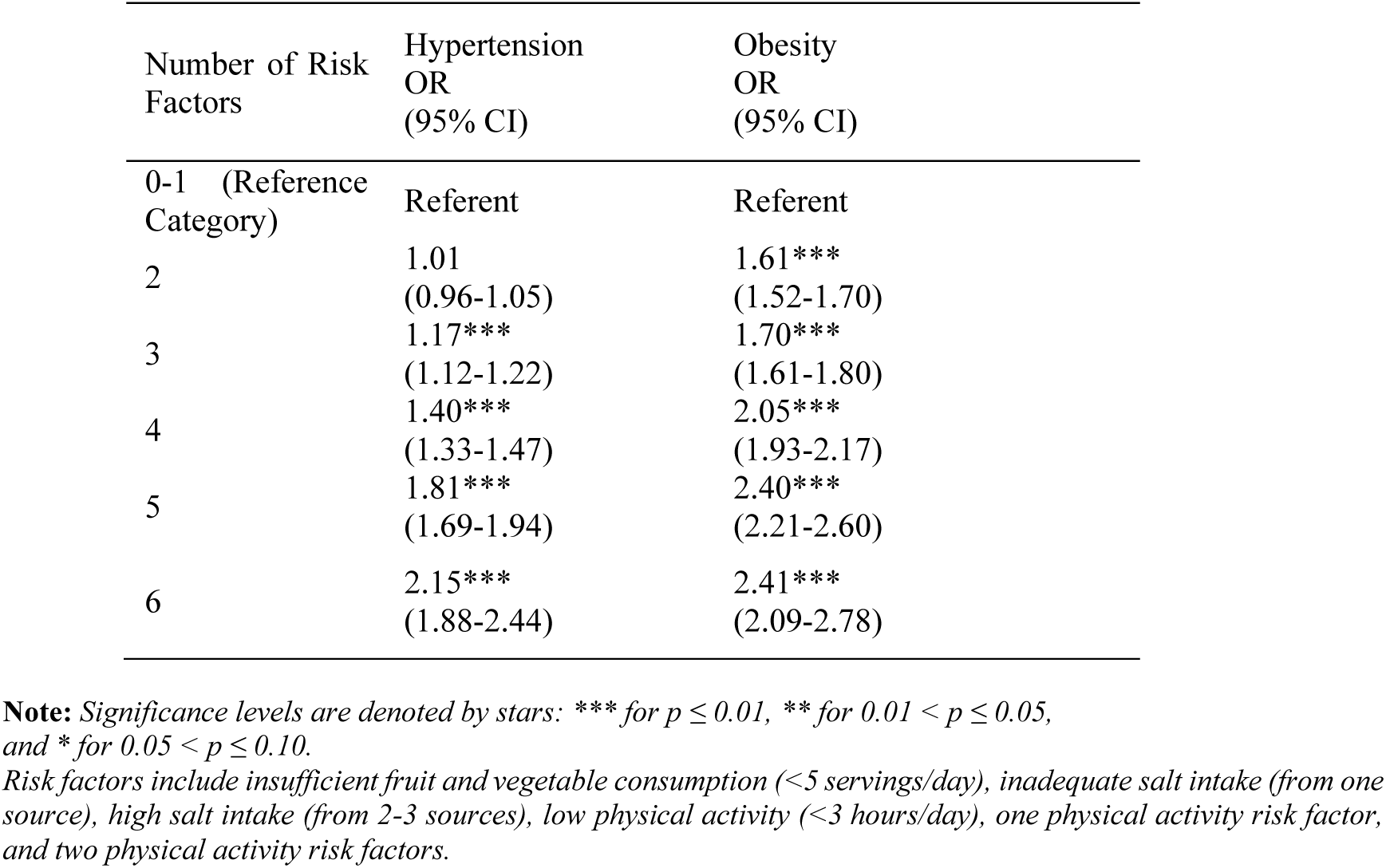
Association between multiple behavioural risk factors and physiological risk factors.

In the multivariable logistic regression models individuals living in urban areas (OR=1.46, 95% CI [1.41-1.51], p<0.001), increasing age (OR for 80+ =3.98, 95% CI [3.70-4.28], p<0.001), consuming fewer than five servings of fruits and vegetables daily (OR=1.52, 95% CI [1.46- 1.60], p<0.001), and having two physical activity risk factors (OR=1.35, 95% CI [1.27-1.43], p<0.001) had significantly higher odds of hypertension (Table 4).

**Table 4.** Adjusted logistic regression model between multiple behavioural risk factors and CVD Physiological Risk Factors.

|  | Hypertension<br>OR (95% CI) | Diabetes<br>OR (95% CI) | Overweight/Obesity<br>OR (95% CI) |
| --- | --- | --- | --- |
| **Location** |  |  |  |
| Rural | Referent | Referent | Referent |
| Urban | 1.46 *** (1.41-1.51) | 1.84 *** (1.66-2.05) | 1.66 *** (1.59-1.72) |
| **Sex** |  |  |  |
| Female | Referent | Referent | Referent |
| Male | 0.84 *** (0.81-0.87) | 0.90 (0.81-1.01) | 0.37 *** (0.36-0.38) |
| **Age Group** |  |  |  |
| 40-49 | Referent | Referent | Referent |
| 50-59 | 2.20 *** (2.02-2.18) | 0.98 *** (0.83-1.16) | 1.01 (0.97-1.06) |
| 60-69 | 3.02 *** (2.88-3.17) | 3.42 *** (2.98-3.93) | 0.99 (0.94-1.04) |
| 70-79 | 3.74 *** (3.53-3.97) | 3.82 *** (3.28-4.45) | 0.55 *** (0.51-0.59) |
| 80+ | 3.98 *** (3.70-4.28) | 1.64 *** (1.31-2.05) | 0.39 *** (0.35-0.43) |
| **Fruit and Vegetables** |  |  |  |
| >5FVeg | Referent | Referent | Referent |
| <5FVeg | 1.52 *** (1.46-1.60) | 1.61 *** (1.36-1.90) | 0.99 (0.94-1.04) |
| **Salt Intake** |  |  |  |
| <1 source of salt intake | Referent | Referent | Referent |
| 2 or 3 sources of salt intake | 0.94 *** (0.91-0.97) | 1.34 *** (1.21-1.49) | 1.21 *** (1.17-1.25) |
| **Physical Activity Risk factors** |  |  |  |
| 0 Physical Activity risk factors | Referent | Referent | Referent |
| 1 Physical Activity risk factors | 1.22 *** (1.18-1.26) | 0.94 (0.84-1.06) | 1.12 *** (1.08-1.16) |
| 2 Physical Activity risk factors | 1.35 *** (1.27-1.43) | 1.47 *** (1.26-1.71) | 1.30 *** (1.22-1.39) |

**Note:** Significance levels are denoted by stars: \*\*\* for $p \leq 0.01$ , \*\* for $0.01 < p \leq 0.05$ , and \* for $0.05 < p \leq 0.10$

Higher odds of diabetes were observed among individuals living in urban areas (OR=1.84, 95% CI [1.66-2.05], p<0.001), increasing age (OR for 70-79 years =3.82, 95% CI [3.28-4.45], p<0.001), consuming fewer than five servings of fruits and vegetables daily (OR=1.61, 95% CI [1.36-1.90], p<0.001), consuming salt from two or three sources (OR=1.34, 95% CI [1.21-1.49], p<0.001), and having two physical activity risk factors (OR=1.47, 95% CI [1.26-1.71], p<0.001).

Urban residents showed higher odds of overweight/obesity (OR=1.66, 95% CI [1.59-1.72], p<0.001), as did those consuming salt from two or three sources (OR=1.21, 95% CI [1.17-1.25], p<0.001) and those with two physical activity risk factors (OR=1.30, 95% CI [1.22-1.39], p<0.001). Conversely, male individuals (OR=0.37, 95% CI [0.36-0.38], p<0.001) and older age groups (70-79 years: OR=0.55, 95% CI [0.51-0.59], p<0.001; 80+ years: OR=0.39, 95% CI [0.35-0.43], p<0.001) had significantly lower odds of overweight/obesity.

## Discussion

This study identified significant associations between various behavioural and physiological risk factors for hypertension, diabetes, and obesity, highlighting important public health implications for Sierra Leone. The findings underscore the necessity of reducing poor dietary habits and increasing physical activity to mitigate the onset of these chronic conditions, which aligns with global and regional health priorities such as Goal 3 of SDGs 2030.

Consistent with existing literature, our results underscore the protective role of adequate fruit and vegetable intake. Participants who consumed fewer than five servings of fruits and vegetables daily were found to have a higher likelihood of experiencing hypertension, diabetes, and overweight/obesity. This pattern highlights the significant health risks associated with inadequate fruit and vegetable intake. This finding aligns with prior studies that have demonstrated the association between higher fruit and vegetable consumption and reduced risk of CVD, likely due to their high content of dietary fibre, vitamins, and minerals such as potassium and magnesium ^28–32^.

Unexpectedly, our study found that higher salt intake was associated with lower odds of hypertension but higher odds of diabetes and overweight/obesity. This finding contradicts established research. ^33^Further research is needed to explore this relationship in the Sierra Leonean context. Salt sensitivity is common among people of African descent, which makes it unlikely that this finding is a causal relationship^34–36^.

The association between physical inactivity and increased risk of diabetes and obesity is well-documented ^37–42^. Our study supports these findings, showing that participants with two physical activity risk factors had higher odds of diabetes and overweight/obesity. This is consistent with studies conducted in West African populations and underscores the need for interventions promoting physical activity to reduce the prevalence of these conditions ^38^ ^43^ ^44^.. The results also align with Sullivan *et al*. ^45^, who reported that US individuals with two physical activity risk factors (sedentary for over three hours daily and low weekly physical activity) had twice the risk of diabetes compared to those with 0-1 physical activity risk factors. Together, this suggests low weekly physical activity and daily sedentariness may play a significant role in the onset and establishment of diabetes. Peoples’ participation in physical activity could be influenced by the built and natural environment in which they live and by personal factors such as gender, age, ability, and motivation. For Sierra Leoneans to be physically active and reduce sedentary time, physical activity programs and interventions must be proposed for urban sector-based jobs while raising awareness of the harmful effects of prolonged sitting.

Participants who consumed 2-3 sources of salt intake showed 1.8 times higher risk of obesity. After adjusting for confounders, such as location, sex and age, the current study found that 2-3 sources of salt intake were associated with a 21% higher risk of obesity. The findings agree with the broader literature that suggest high salt intake is associated with obesity ^46–49^. In the case of Sierra Leone, it is likely that the drivers behind higher salt intake and obesity could be explained by increased consumption of processed calorie-dense foods, which can lead to excessive sodium intake ^46–50^.

Our study contributes valuable insights into the epidemiology of CVD risk factors in sub-Saharan Africa, particularly in Sierra Leone. The findings suggest that urbanization and associated behaviour changes, such as consumption of processed foods and low physical activity, are contributing to the rising burden of CVDs in this region. Given the high prevalence of hypertension, diabetes, and obesity found in this study, targeted public health strategies are urgently needed to address these risk factors.

For Sierra Leone, specific interventions could include public health campaigns to raise awareness about the benefits of fruit and vegetable consumption and the risks associated with high salt intake. Policies to promote physical activity, such as creating exercise facilities and organizing community fitness events, could also be beneficial. Additionally, further research is needed to explore the complex interplay between behavioural, environmental, and genetic factors in the development of CVDs in this context.

### Limitations of Current Research

This study has several limitations that must be acknowledged. First, the cross-sectional design limits our ability to infer causality, as it captures data at a single point in time and prevents establishing temporal relationships between exposure and outcome. Therefore, while associations can be identified, causation cannot be definitively determined.

Second, reliance on self-reported dietary and physical activity data introduces potential recall bias and measurement errors. Participants might not accurately remember or report their dietary intake and physical activity levels, leading to misclassification and potential bias in the findings. Although efforts were made to minimize these errors, such as using standardized questionnaires and trained data collectors, the inherent limitations of self-report data remain. Third, the study was conducted exclusively among Sierra Leonean adults aged 40 years and above. This age restriction means that the findings may not be generalizable to younger populations or to other regions with different demographic profiles. The focus on an older population was driven by the higher prevalence of CVD risk factors in this age group, but future studies should include a broader age range to enhance generalizability.

Additionally, the study did not account for all possible confounding variables. While key sociodemographic factors were adjusted for, other potential confounders, such as genetic predispositions, environmental exposures, and access to healthcare, were not included. These unmeasured confounders might have influenced the observed associations and should be considered in future research.

Finally, despite efforts to ensure accuracy, the possibility of dietary assessment measurement error by participants exists. Despite these limitations, the study provides important insights into the behavioural and physiological risk factors for CVDs in Sierra Leone and highlights areas for future research and public health interventions.

## Conclusion

In conclusion, this study reveals significant associations between dietary habits, physical activity levels, and three physiological risk factors for CVD; hypertension, diabetes, and obesity among adults in Bo District, Sierra Leone. The findings emphasize the critical need for targeted public health interventions aimed at promoting healthier dietary practices and increasing physical activity to mitigate the growing burden of CVDs in this region.

Addressing these modifiable risk factors through public health campaigns, policy changes, and community-based interventions could significantly reduce the prevalence of CVDs and improve overall population health. Additionally, the study highlights the importance of further research to explore the complex interplay between behavioural, environmental, and genetic factors in the development of CVDs.

By understanding and addressing the specific risk factors prevalent in Sierra Leone, effective strategies can be developed to combat CVDs and contribute to the global efforts to reduce the burden of non-communicable diseases. These efforts are crucial not only for improving health outcomes but also for achieving sustainable development goals related to health and well-being in low- and middle-income countries.

## Data Availability

All data produced in the present study are available upon reasonable request to the authors

## Appendix

### Prevalence of CVD Risk Factors by individual Behavioural factors

Participants who consumed fewer than five servings of fruits and vegetables daily exhibited higher prevalence of hypertension (85.8% vs. 14.2%), diabetes (89% vs. 10.4%), and overweight/obesity (84.7% vs. 15.3%) compared with those who met the recommended intake of five servings per day (Table 1).

In contrast, participants with higher salt intake, defined as consuming salt from two or three sources, demonstrated a lower prevalence of hypertension (46.1% vs. 53.9%) and overweight/obesity (47.1% vs. 52.9%) but a higher prevalence of diabetes (51.1% vs. 42.9%) compared with those with lower salt intake.

Physical activity also had a significant association with the prevalence of physiological CVD risk factors. Individuals with at least one physical activity risk factor (e.g., inadequate moderate and vigorous physical activity (MVPA) or high levels of physical inactivity) showed higher prevalence rates of hypertension (42.0% vs. 40.5%), diabetes (40.8% vs. 38.9%), and overweight/obesity (44.4% vs. 41.8%) compared with those with no physical activity risk factors (Table 1).

**Table 1:**
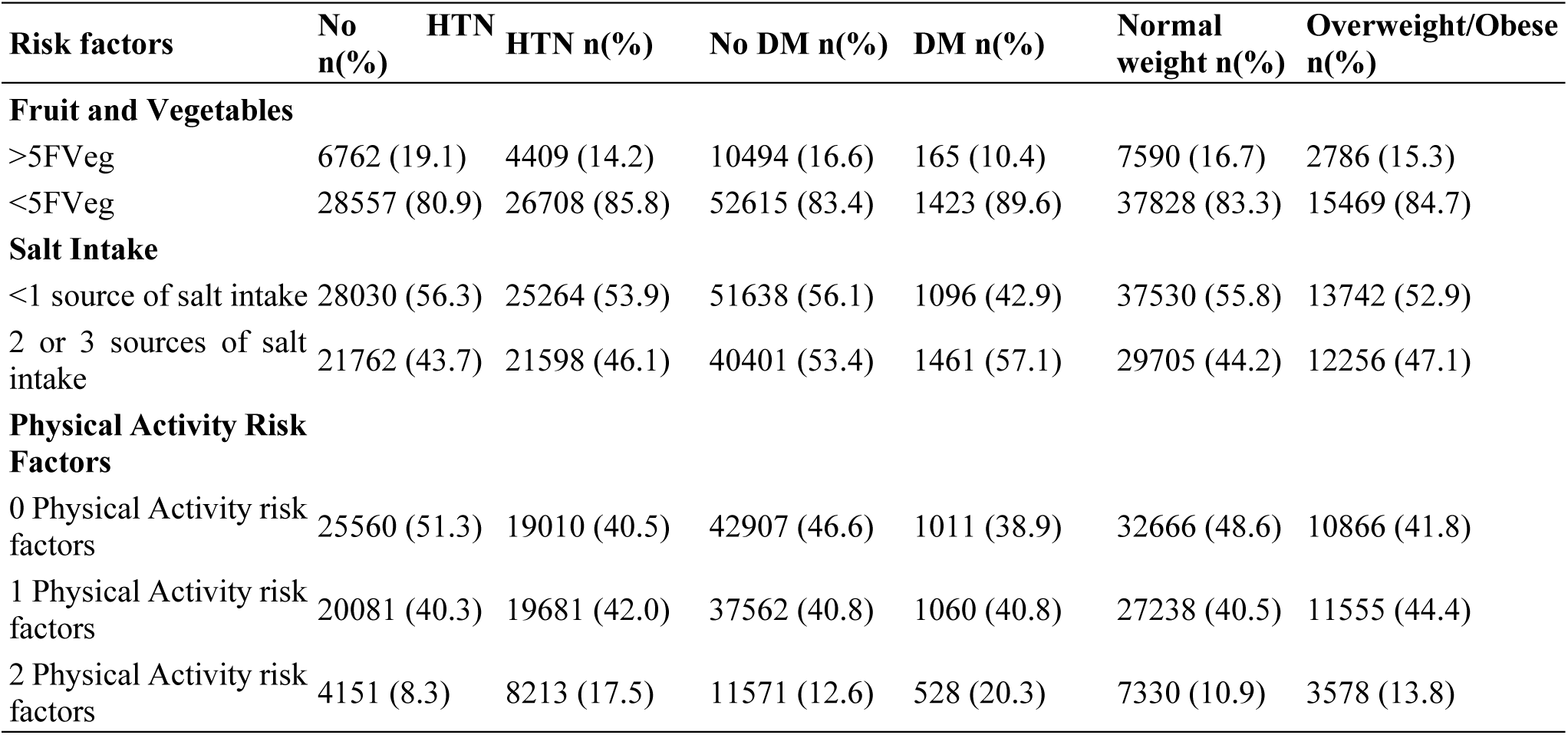
Descriptive Analysis of Behavioural Risk Factors and CVD Physiological Risk Factors.

Sociodemographic characteristics were associated with the prevalence of physiological CVD risk factors. Rural residents had a higher prevalence of diabetes and overweight/obesity but a lower prevalence of hypertension compared with urban residents (Table 5). This variation may reflect differences in behaviour and access to healthcare services between rural and urban areas.

Sex differences were also apparent, with males exhibiting lower prevalence rates of hypertension (46.2% vs. 53.8%), diabetes (45.9% vs. 54.1%), and overweight/obesity (33.3% vs. 66.7%) compared with females. Age-wise, individuals aged 40-49 years had higher prevalence rates of hypertension (32.4%), diabetes (31.1%), and overweight/obesity (47.9%) compared to older age groups (Table 2). This suggests that middle-aged adults in this cohort may be at greater risk and highlights the need for targeted interventions.

**Table 2:**
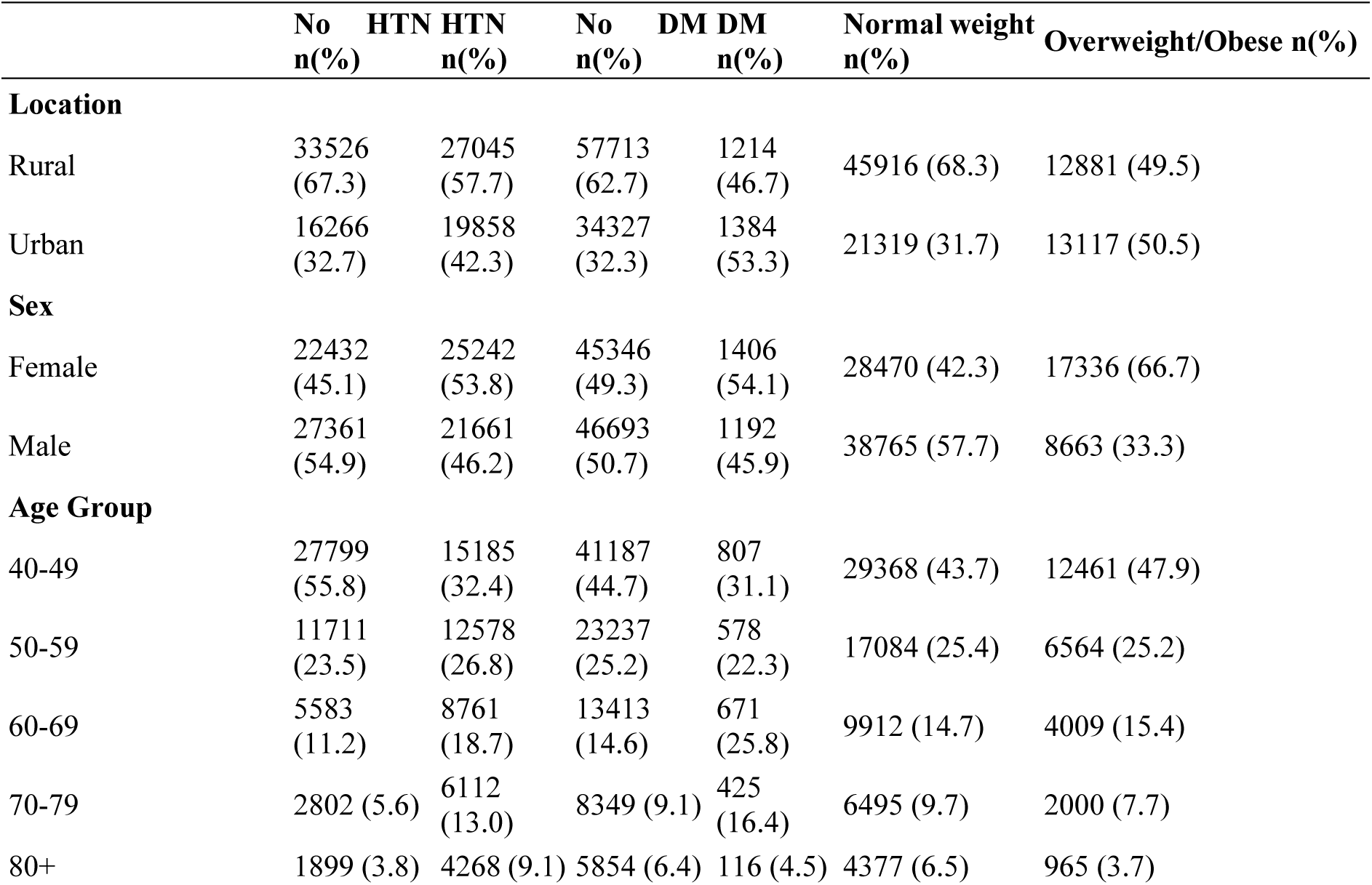
Descriptive Analysis of Sociodemographic Characteristics by Hypertension, Diabetes, and Weight Status.

### Association Between Behavioural Risk Factors, Sociodemographic Characteristics, and Physiological Risk Factors for CVD

The results from the unadjusted regression analysis reveal that individuals consuming fewer than five servings of fruits and vegetables daily had significantly higher odds of hypertension (OR=1.62, 95% CI [1.56-1.69], p<0.001), diabetes (OR=1.82, 95% CI [1.54-2.15], p<0.001), and overweight/obesity (OR=1.17, 95% CI [1.12-1.23], p<0.001) compared with those meeting the recommended intake (Table 3).

**Table 3:** Unadjusted Logistic Regression Models for Behavioural Risk Factors and Cardiovascular Conditions.

| Risk Factors | Hypertension<br>OR<br>(95% CI) | Diabetes<br>OR<br>(95% CI) | Overweight/Obesity<br>OR<br>(95% CI) |
| --- | --- | --- | --- |
| **Fruit and Vegetables** |  |  |  |
| >5FVeg | Referent | Referent | Referent |
| <5FVeg | 1.62*** (1.56-1.69) | 1.82*** (1.54-2.15) | 1.17*** (1.12-1.23) |
| **Salt Intake** |  |  |  |
| <1 source of salt intake | Referent | Referent | Referent |
| 2 or 3 sources of salt intake | 0.94*** (0.91-0.97) | 1.36*** (1.23-1.51) | 1.18*** (1.14-1.22) |
| **Physical Activity Risk factors** |  |  |  |
| 0 Physical Activity risk factors | Referent | Referent | Referent |
| 1 Physical Activity risk factors | 1.40*** (1.35-1.45) | 1.20*** (1.07-1.35) | 1.22*** (1.17-1.26) |
| 2 Physical Activity risk factors | 2.40*** (2.27-2.52) | 2.31*** (2.01-2.65) | 1.26*** (1.19-1.34) |

**Note:** Significance levels are denoted by stars: \*\*\* for $p \leq 0.01$ , \*\* for $0.01 < p \leq 0.05$ , and \* for $0.05 < p \leq 0.10$

In terms of salt intake, participants consuming salt from two or three sources had higher odds of diabetes (OR=1.36, 95% CI [1.23-1.51], p<0.001) and overweight/obesity (OR=1.18, 95% CI [1.14-1.22], p<0.001), but lower odds of hypertension (OR=0.94, 95% CI [0.91-0.97], p<0.001) compared with those with lower salt intake. For physical activity, individuals with two physical activity risk factors had significantly higher odds of hypertension (OR=2.40, 95% CI [2.27-2.52], p<0.001), diabetes (OR=2.31, 95% CI [2.01-2.65], p<0.001), and overweight/obesity (OR=1.26, 95% CI [1.19-1.34], p<0.001) compared with those with no physical activity risk factors (Table 3).

## References

1. Chand SS, Singh B, Kumar S. The economic burden of non-communicable disease mortality in the South Pacific: Evidence from Fiji. PLOS ONE 2020;15(7):e0236068. doi: 10.1371/journal.pone.0236068

2. Gowshall M, Taylor-Robinson SD. The increasing prevalence of non-communicable diseases in low-middle income countries: the view from Malawi. International Journal of General Medicine 2018;11(null):255–64. doi: 10.2147/IJGM.S157987

3. UN. United Nations-Department of Economic and Social Affairs-Sustainable Development Goal 3: Ensure healthy lives and promote well-being for all at all ages 2015 [cited 2025 5 September]. Available from: https://sdgs.un.org/goals/goal32024

4. Vos T, Lim SS, Abbafati C, et al. Global burden of 369 diseases and injuries in 204 countries and territories, 1990&#x2013;2019: a systematic analysis for the Global Burden of Disease Study 2019. The Lancet 2020;396(10258):1204–22. doi: 10.1016/S0140-6736(20)30925-9

5. Amini M, Zayeri F, Salehi M. Trend analysis of cardiovascular disease mortality, incidence, and mortality-to-incidence ratio: results from global burden of disease study 2017. BMC Public Health 2021;21(1):401. doi: 10.1186/s12889-021-10429-0

6. Roth GA, Mensah GA, Johnson CO, et al. Global Burden of Cardiovascular Diseases and Risk Factors, 1990–2019: Update From the GBD 2019 Study. Journal of the American College of Cardiology 2020;76(25):2982–3021. doi: 10.1016/j.jacc.2020.11.010

7. McKenzie BL, Santos JA, Geldsetzer P, et al. Evaluation of sex differences in dietary behaviours and their relationship with cardiovascular risk factors: a cross-sectional study of nationally representative surveys in seven low- and middle-income countries. Nutrition Journal 2020;19(1):3. doi: 10.1186/s12937-019-0517-4

8. Rosengren A, Smyth A, Rangarajan S, et al. Socioeconomic status and risk of cardiovascular disease in 20 low-income, middle-income, and high-income countries: the Prospective Urban Rural Epidemiologic (PURE) study. The Lancet Global Health 2019;7(6):e748–e60. doi: 10.1016/S2214-109X(19)30045-2

9. Yusuf S, Joseph P, Rangarajan S, et al. Modifiable risk factors, cardiovascular disease, and mortality in 155&#x2008;722 individuals from 21 high-income, middle-income, and low-income countries (PURE): a prospective cohort study. The Lancet 2020;395(10226):795–808. doi: 10.1016/S0140-6736(19)32008-2

10. de Oliveira Otto MC, Afshin A, Micha R, et al. The Impact of Dietary and Metabolic Risk Factors on Cardiovascular Diseases and Type 2 Diabetes Mortality in Brazil. PLOS ONE 2016;11(3):e0151503. doi: 10.1371/journal.pone.0151503

11. WHO. World Health Organization Noncommunicable Disease Surveillance, Monitoring and Reporting - STEPS surveillance manual. STEPs manual. 2017 [updated 26 January 2017. Available from: https://www.who.int/teams/noncommunicable-diseases/surveillance/systems-tools/steps/manuals accessed September 2024.

12. Gafane-Matemane LF, Craig A, Kruger R, et al. Hypertension in sub-Saharan Africa: the current profile, recent advances, gaps, and priorities. Journal of Human Hypertension 2024 doi: 10.1038/s41371-024-00913-6

13. Green R, Cornelsen L, Dangour AD, et al. The effect of rising food prices on food consumption: systematic review with meta-regression. BMJ: British Medical Journal 2013;346:f3703. doi: 10.1136/bmj.f3703

14. Popkin BM, Adair LS, Ng SW. Global nutrition transition and the pandemic of obesity in developing countries. Nutrition Reviews 2012;70(1):3–21. doi: 10.1111/j.1753-4887.2011.00456.x

15. Smart JC, Tschirley D, Smart F. Diet Quality and Urbanization in Mozambique. Food and Nutrition Bulletin 2020;41(3):298–317. doi: 10.1177/0379572120930123

16. Steyn N. Dietary changes and the health transition in South Africa: implications for health policy NP Steyn, D. Bradshaw, R. Norman, JD Joubert, M. Schneider and K. Steyn. The double burden of malnutrition: Case Studies from six developing countries 2006;84:259.

17. Dehghan M, Mente A, Zhang X, et al. Associations of fats and carbohydrate intake with cardiovascular disease and mortality in 18 countries from five continents (PURE): a prospective cohort study. The Lancet 2017;390(10107):2050–62. doi: 10.1016/S0140-6736(17)32252-3

18. Jones-Smith JC, Gordon-Larsen P, Siddiqi A, et al. Is the burden of overweight shifting to the poor across the globe? Time trends among women in 39 low- and middle-income countries (1991–2008). International Journal of Obesity 2012;36(8):1114–20. doi: 10.1038/ijo.2011.179

19. Muthuri SK, Francis CE, Wachira L-JM, et al. Evidence of an Overweight/Obesity Transition among School-Aged Children and Youth in Sub-Saharan Africa: A Systematic Review. PLOS ONE 2014;9(3):e92846. doi: 10.1371/journal.pone.0092846

20. Oyebode O, Oti S, Chen Y-F, et al. Salt intakes in sub-Saharan Africa: a systematic review and meta-regression. Population Health Metrics 2016;14(1):1. doi: 10.1186/s12963-015-0068-7

21. Ziraba AK, Fotso JC, Ochako R. Overweight and obesity in urban Africa: A problem of the rich or the poor? BMC Public Health 2009;9(1):465. doi: 10.1186/1471-2458-9-465

22. Mensah DO, Nunes AR, Bockarie T, et al. Meat, fruit, and vegetable consumption in sub-Saharan Africa: a systematic review and meta-regression analysis. Nutrition Reviews 2020;79(6):651–92. doi: 10.1093/nutrit/nuaa032

23. Addo J, Smeeth L, Leon DA. Hypertension In Sub-Saharan Africa. Hypertension 2007;50(6):1012–18. doi: 10.1161/HYPERTENSIONAHA.107.093336

24. Ataklte F, Erqou S, Kaptoge S, et al. Burden of Undiagnosed Hypertension in Sub-Saharan Africa. Hypertension 2015;65(2):291–98. doi: 10.1161/HYPERTENSIONAHA.114.04394

25. Atibila F, Hoor Gt, Donkoh ET, et al. Prevalence of hypertension in Ghanaian society: a systematic review, meta-analysis, and GRADE assessment. Systematic Reviews 2021;10(1):220. doi: 10.1186/s13643-021-01770-x

26. Bosu WK, Bosu DK. Prevalence, awareness and control of hypertension in Ghana: A systematic review and meta-analysis. PLOS ONE 2021;16(3):e0248137. doi: 10.1371/journal.pone.0248137

27. Stats SL. Sierra Leone population and housing census 2015; Statistics Sierra Leone; 2015 [cited 2024. Available from: https://www.statistics.sl/index.php/census/census-2015.html accessed 17 February 2024.

28. Aune D, Giovannucci E, Boffetta P, et al. Fruit and vegetable intake and the risk of cardiovascular disease, total cancer and all-cause mortality—a systematic review and dose-response meta-analysis of prospective studies. International Journal of Epidemiology 2017;46(3):1029–56. doi: 10.1093/ije/dyw319

29. Oyebode O, Gordon-Dseagu V, Walker A, et al. Fruit and vegetable consumption and all-cause, cancer and CVD mortality: analysis of Health Survey for England data. Journal of Epidemiology and Community Health 2014;68(9):856. doi: 10.1136/jech-2013-203500

30. Wallace TC, Bailey RL, Blumberg JB, et al. Fruits, vegetables, and health: A comprehensive narrative, umbrella review of the science and recommendations for enhanced public policy to improve intake. Critical Reviews in Food Science and Nutrition 2020;60(13):2174–211. doi: 10.1080/10408398.2019.1632258

31. Zhan J, Liu Y-J, Cai L-B, et al. Fruit and vegetable consumption and risk of cardiovascular disease: A meta-analysis of prospective cohort studies. Critical Reviews in Food Science and Nutrition 2017;57(8):1650–63. doi: 10.1080/10408398.2015.1008980

32. Zurbau A, Au-Yeung F, Blanco Mejia S, et al. Relation of Different Fruit and Vegetable Sources With Incident Cardiovascular Outcomes: A Systematic Review and Meta-Analysis of Prospective Cohort Studies. Journal of the American Heart Association 2020;9(19):e017728. doi: 10.1161/JAHA.120.017728

33. Islam SMS, Mainuddin A, Islam MS, et al. Prevalence of risk factors for hypertension: A cross-sectional study in an urban area of Bangladesh. Global Cardiology Science and Practice 2015;2015(4) doi: 10.5339/gcsp.2015.43

34. Aburto NJ, Ziolkovska A, Hooper L, et al. Effect of lower sodium intake on health: systematic review and meta-analyses. Bmj 2013;346:f1326.

35. Appel LJ, Moore TJ, Obarzanek E, et al. A Clinical Trial of the Effects of Dietary Patterns on Blood Pressure. New England Journal of Medicine 1997;336(16):1117–24. doi: doi:10.1056/NEJM199704173361601

36. He FJ, Li J, MacGregor GA. Effect of longer term modest salt reduction on blood pressure: Cochrane systematic review and meta-analysis of randomised trials. BMJ: British Medical Journal 2013;346:f1325. doi: 10.1136/bmj.f1325

37. Aune D, Norat T, Leitzmann M, et al. Physical activity and the risk of type 2 diabetes: a systematic review and dose–response meta-analysis. European Journal of Epidemiology 2015;30(7):529–42. doi: 10.1007/s10654-015-0056-z

38. Cleven L, Krell-Roesch J, Nigg CR, et al. The association between physical activity with incident obesity, coronary heart disease, diabetes and hypertension in adults: a systematic review of longitudinal studies published after 2012. BMC Public Health 2020;20(1):726. doi: 10.1186/s12889-020-08715-4

39. Jakicic JM, Kraus WE, Powell KE, et al. Association between Bout Duration of Physical Activity and Health: Systematic Review. Med Sci Sports Exerc 2019;51(6):1213–19. doi: 10.1249/mss.0000000000001933

40. Jeon CY, Lokken RP, Hu FB, et al. Physical Activity of Moderate Intensity and Risk of Type 2 Diabetes: A systematic review. Diabetes Care 2007;30(3):744–52. doi: 10.2337/dc06-1842

41. Rauner A, Mess F, Woll A. The relationship between physical activity, physical fitness and overweight in adolescents: a systematic review of studies published in or after 2000. BMC Pediatrics 2013;13(1):19. doi: 10.1186/1471-2431-13-19

42. Smith AD, Crippa A, Woodcock J, et al. Physical activity and incident type 2 diabetes mellitus: a systematic review and dose–response meta-analysis of prospective cohort studies. Diabetologia 2016;59(12):2527–45. doi: 10.1007/s00125-016-4079-0

43. Huang Z, Zhuang X, Huang R, et al. Physical Activity and Weight Loss Among Adults With Type 2 Diabetes and Overweight or Obesity: A Post Hoc Analysis of the Look AHEAD Trial. JAMA Network Open 2024;7(2):e240219–e19. doi: 10.1001/jamanetworkopen.2024.0219

44. Lee IM, Shiroma EJ, Lobelo F, et al. Effect of physical inactivity on major non-communicable diseases worldwide: an analysis of burden of disease and life expectancy. The Lancet 2012;380(9838):219-29. doi: 10.1016/S0140-6736(12)61031-9

45. Sullivan PW, Morrato EH, Ghushchyan V, et al. Obesity, Inactivity, and the Prevalence of Diabetes and Diabetes-Related Cardiovascular Comorbidities in the U.S., 2000–2002. Diabetes Care 2005;28(7):1599–603. doi: 10.2337/diacare.28.7.1599

46. Grimes CA, Bolhuis DP, He FJ, et al. Dietary sodium intake and overweight and obesity in children and adults: a protocol for a systematic review and meta-analysis. Syst Rev 2016;5:7. doi: 10.1186/s13643-015-0175-3 [published Online First: 20160118]

47. Lee J, Sohn C, Kim OY, et al. The association between dietary sodium intake and obesity in adults by sodium intake assessment methods: a review of systematic reviews and re-meta-analysis. Nutr Res Pract 2023;17(2):175–91. doi: 10.4162/nrp.2023.17.2.175 [published Online First: 20230120]

48. Ma Y, He FJ, MacGregor GA. High Salt Intake. Hypertension 2015;66(4):843–49. doi: 10.1161/HYPERTENSIONAHA.115.05948

49. Moosavian SP, Haghighatdoost F, Surkan PJ, et al. Salt and obesity: a systematic review and meta-analysis of observational studies. International Journal of Food Sciences and Nutrition 2017;68(3):265–77. doi: 10.1080/09637486.2016.1239700

50. MoH, MAFFS. Ministry of Health and Sanitation (MOHS) - SIERRA LEONE FOOD BASED DIETARY GUIDELINE FOR HEALTHY EATING (2016). Food and Agriculture Organization of the United Nations (FAO): Ministry of Agriculture, Forestry and Food Security (MAFFS)-Sierra Leone, 2016.

